# Beta blockers, digoxin or both following an incident diagnosis of atrial fibrillation – a prospective cohort study

**DOI:** 10.1101/2023.05.18.23290189

**Authors:** James M Brophy, Lyne Nadeau

**Author notes:** Corresponding author *Email address:* (James M Brophy MD PhD).

## Abstract

**Background:** Atrial fibrillation is one of the most common arrhythmias but the optimal drug choice for a rate control strategy remains uncertain. In particular, controversy and uncertainty exists regarding the safety of digoxin in this context.

**Methods:** This was a retrospective cohort claims database study of patients with an incident hospital discharge diagnosis of atrial fibrillation between 2011 and 2015. The exposure variables were a discharge prescription for beta blockers, digoxin or both. The primary outcome was a composite of total in-hospital mortality or a repeat cardiovascular (CV) hospitalization. Secondary outcomes were the individual components of the primary outcome. Baseline confounding was controlled with propensity score inverse probability weighting using a entropy balancing algorithm and the prespecified estimand was the average treatment effect among the treated. In sensitivity analyses, baseline covariate imbalances were adjusted using a maximum likelihood algorithm and an overall average treatment effect estimand. Treatment effects for the weighted samples were calculated from a Cox proportional hazards model.

**Results:** 12,723 patients were discharged on beta blockers alone, 406 on digoxin alone, and 1,499 discharged on combined beta blocker / digoxin therapy with a median follow-up time of 356 days. In the unadjusted analyses, the primary outcome occured most frequently in the combined exposure group (15.5%) compared to the isolated digoxin (13.3%) and beta blocker (11.5%) groups (p < 0.001 for trend). There were more CV hospitalizations in the combined beta blocker / digoxin group (14.4%) compared to the BB (10.7%) or digoxin (10.6%) groups (p = 0.006 for trend). There were more deaths in the digoxin group (2.7%) and the combined group (1.1%) groups compared to the BB alone group (0.8%) (p < 0.001 for trend). However, after baseline covariate adjustment, the digoxin alone (hazard ratio (HR) 1.24, 95% CI 0.85 - 1.81) and the combined group (HR 1.09, 95% CI 0.90 - 1.31) were not associated with increased risk for the composite endpoint compared with the beta blocker alone group. These results were robust to sensitivity analyses.

**Conclusion:** After accounting for baseline imbalances, patients hospitalized for incident atrial fibrillation and discharged on digoxin alone or the combination of digoxin and a beta blocker were not associated with an increase in the composite outcome of recurrent CV hospitalizations and death compared to those discharged on isolated beta blocker therapy. However, additional studies are required to refine the precision of these estimates.

## 1. Introduction

Atrial fibrillation is one of the most common arrhythmias and a progressive worldwide increase in its incidence, prevalence, and burden of disease has been observed in the 21st century [5] The rising incidence is likely multifactorial due to increasing longevity with an associated co-existence of both cardiac and non-cardiac comorbidities as well as improved detection strategies, including the use of wearables. Atrial fibrillation may exist in paroxysmal, persistent or chronic forms. Although some atrial fibrillation patients are asymptomatic, many do present to the hospital with symptoms of palpitations, dyspnea or reduced exercise tolerance. Atrial fibrillation frequently co-exists as either a cause or an effect of heart failure and major complications may include systemic or cerebral thromboembolism as well as an increase in overall mortality [1].

There are numerous therapeutic options for the treatment of acute atrial fibrillation including electrical cardioversion and more recently pulmonary vein ablation, but the great majority of patients still receive some pharmacological treatment either as part of a rhythm or rate control strategy. Although there are suggestions that in special situations a rhythm control strategy may offer added value[10], many patients are treated with a rate control strategy that most often includes beta blockers as first line therapy. Numerous randomized trials have shown beta-blockers have a major impact on mortality in individuals with congestive heart failure[4], but there is little evidence to support their selection in patients with atrial fibrillation, even among those with with coexisting heart failure[11]. Digoxin is generally viewed as a second line therapeutic option for atrial fibrillation patients achieving suboptimal rate control with beta blockers and for those with associated left ventricular dysfunction[2]. However controversy remains about digoxin’s adjunctive role in these conditions. For example, a recent meta-analysis of 28 trials[15] to assess the benefits and harms of digoxin for atrial fibrillation and atrial flutter concluded that many suffered from important biases and a severe lack of power to evaluate the clinical effects of digoxin on all-cause mortality and repeat hospitalizations. This observational study seeks to provide further evidence regarding the efficacy and safety of digoxin in patients with incident atrial fibrillation who are treated, or not, with beta blockers.

## 2. Methods

### Design and Population

This retrospective claims database study was conducted using the Truven Health Analytics MarketScan Research Databases with U.S. commercial and Medicare supplemental claims. The dataset included all inpatient and outpatient medical claims, plus enrollment information, within a 5 year period (2011-2015), on individuals with a hospitalization for atrial fibrillation (inpatient medical claim with primary diagnosis using International Classification of Diseases, Ninth Revision, Clinical Modification [ICD-9-CM] code 427.x or ICD-10 diagnosis codes I46, I47, I 48, I49, I51.8, R00.1, R00.8, R01.2). Subjects had to be continuously enrolled for at least 12 months before the index date with no prior atrial fibrillation diagnosis. The discharge date of the first observed hospital claim for atrial fibrillation was the assigned index date for this incident cohort. The cohort was restricted to patients who received within 2 days of discharge a prescription for a beta blocker, excluding sotalol, digoxin or both. Specifically, patients receiving concomitant therapy with other anti-arrhythmic drug therapies including calcium channel blockers, amiodarone, dronedarone, flecainide, propafenone, dofetilide, mexiletine or disopyramide were excluded. The alignment of study eligibility, treatment assignment, and follow-up mininizes the possibility of prevalent user, selection, misclassification and immortal time biases[9]. The primary outcome was a repeat cardiovascular hospitalization or within hospital death occurring after the initial hospital discharge date. The project received ethical approved from the McGill University Health Center research ethics board.

### Propensity score methods

For observational study designs, weighting by propensity scores (the probability of being assigned to a certain treatment conditional on baseline characteristics and estimated using logistic regression) is an effective strategy for achieving good baseline balance and reducing or eliminating parametric model dependencies[3]. Weighting subjects by the inverse probability of treatment received creates a synthetic sample in which treatment assignment becomes independent of measured baseline covariates. This pre-processing by propensity scores, by balancing the two groups, allows one to obtain unbiased estimates of average treatment effects. Essentially this means for any given propensity score, the choice of exposure group is a random process, at least as far as the measured confounders are concerned. In a randomized clinical trial, by nature of the trial design the whole study population is eligible for treatment and the most common effect measure is the average treatment effect **(ATE)**. However in a real world population study with observational data, the average treatment effect of the treated **(ATT)**, is often the more appropriate effect measure as it avoids the medically or scientifically questionable assumption that every participant can be switched from their current treatment to the opposite. **ATT** is a relevant estimand because interest is generally in the causal effect among patients eligible or selected for a particular treatment. Therefore, we used an **ATT** estimand in our primary analyses and an **ATE** estimand in our sensitivity analyses.

### Statistical analyses

The Cochran Armitage (Mantel-Haenszel *X*^2^) test for trend was used to evaluate the crude binomial outcome proportions across the different exposure levels. A multi-category propensity treatment score was created by including all available baseline variables in a multivariate logistic regression model[7]. We used inverse probability weights using a entropy balancing algorithm from the propensity scores generated from a logistic regression model thereby creating pseudo populations with balance between the covariate profiles of the different exposures groups. The success of the propensity score methods were assessed by examining standardized mean difference between the groups and by graphical inspection of the propensity score histograms[6]. In the primary analysis, the adjusted effect of group exposure on the time to the composite outcome was modeled with Cox proportional hazards regression using the weighted datasets with robust standard errors. We also calculated the Evalue[17], defined as the minimum strength of association on the risk-ratio scale that an unmeasured confounder would need to have with both the treatment assignment and the outcome to reverse a specific treatment-outcome association, conditional on the measured covariates.

All analyses were performed in R[16], specifically using the WeightIt package [7] to create the pseudo populations and the Cobalt[6] package to assess the success in achieving covariate balance. Evalues were calculated using the EValue package[17] in R.

## 3. Results

During the study period, among a total of 52,164 hospitalizations for atrial fibrillation, there were 12,723 patients discharged on beta blockers alone, 406 on digoxin alone, and 1,499 discharged on combined beta blocker / digoxin therapy. Table 1 compares their baseline characteristics. In general, the exposure groups with digoxin were older (p < 0.001) and had more comorbidites as assessed by the higher Charlson index score (p < 0.001) and more cancer diagnoses (p < 0.001) compared to the beta blocker alone group. As expected, the digoxin groups also had more congestive heart failure and chronic obstructive pulmonary diagnoses (both p < 0.001). The overall low rate of baseline anticoagulants is to be expected as this was an incident cohort of new atrial fibrillation cases. The higher discharge rate of anticoagulation in the beta blocker / digoxin group is also expected given the higher CHADS-VASC score in this group compared to the other groups (p < 0.001). The digoxin alone group also had fewer in-hospital cardioversions (p < 0.007). As pre-specified in the protocol, none of these three exposure groups received other anti-arrhythmic drugs on discharge.

**Table 1:**
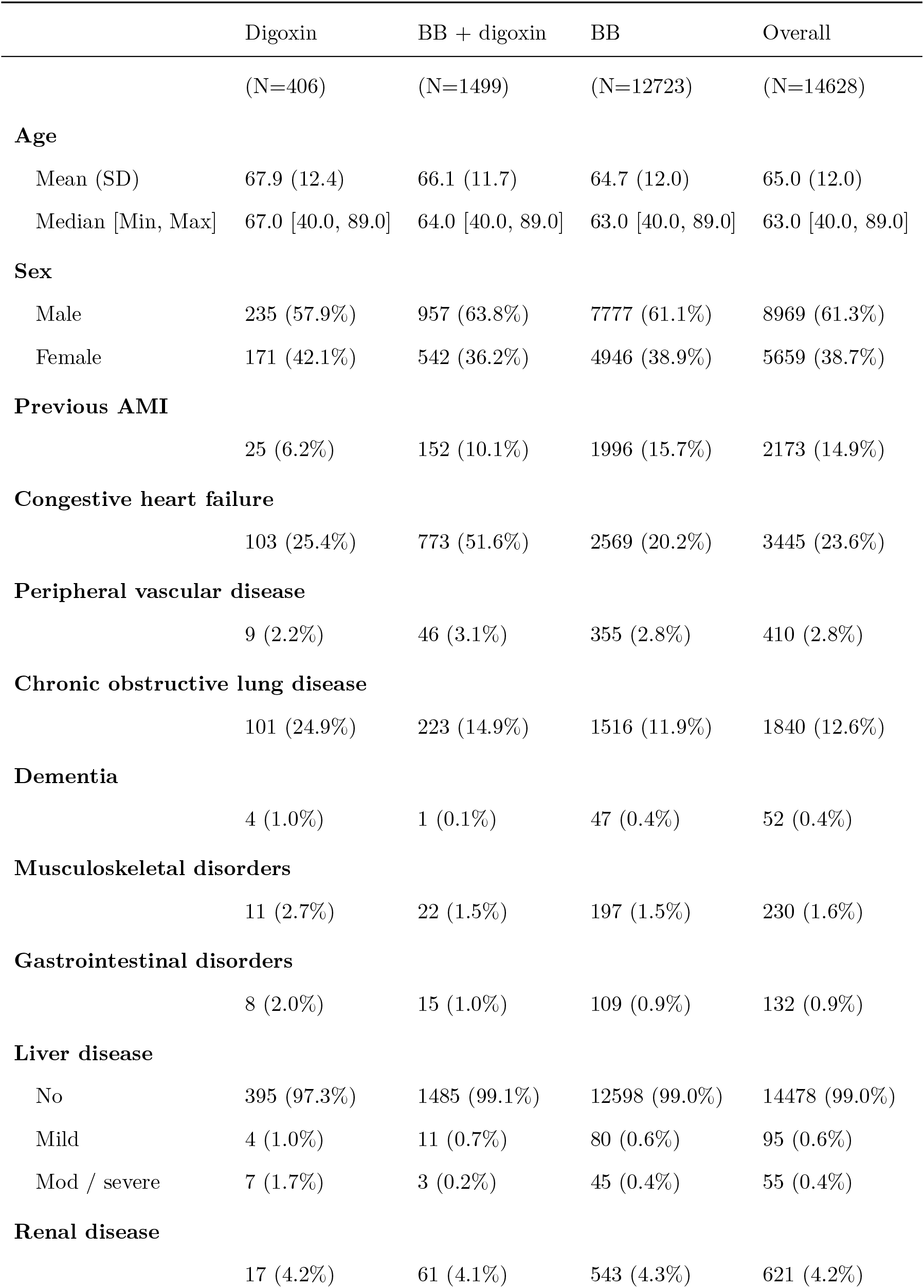
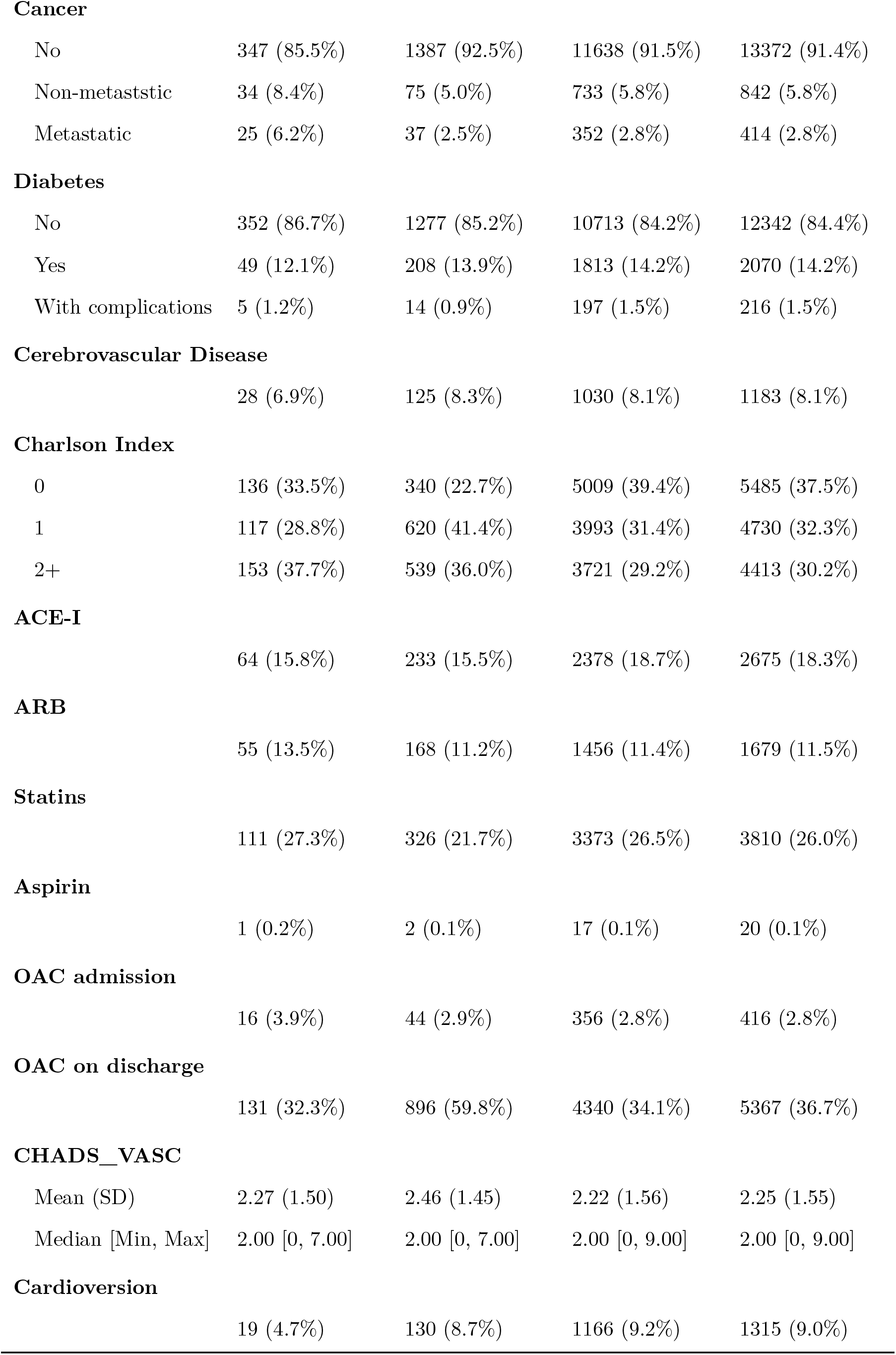
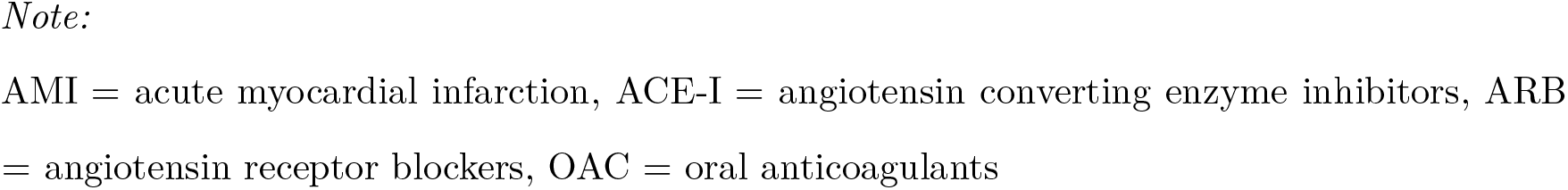
Table 1 Demographic and Baseline Characteristics

|  | Digoxin | BB + digoxin | BB | Overall |
| --- | --- | --- | --- | --- |
|  | (N=406) | (N=1499) | (N=12723) | (N=14628) |
| **Age** |  |  |  |  |
| Mean (SD) | 67.9 (12.4) | 66.1 (11.7) | 64.7 (12.0) | 65.0 (12.0) |
| Median [Min, Max] | 67.0 [40.0, 89.0] | 64.0 [40.0, 89.0] | 63.0 [40.0, 89.0] | 63.0 [40.0, 89.0] |
| **Sex** |  |  |  |  |
| Male | 235 (57.9%) | 957 (63.8%) | 7777 (61.1%) | 8969 (61.3%) |
| Female | 171 (42.1%) | 542 (36.2%) | 4946 (38.9%) | 5659 (38.7%) |
| **Previous AMI** |  |  |  |  |
|  | 25 (6.2%) | 152 (10.1%) | 1996 (15.7%) | 2173 (14.9%) |
| **Congestive heart failure** |  |  |  |  |
|  | 103 (25.4%) | 773 (51.6%) | 2569 (20.2%) | 3445 (23.6%) |
| **Peripheral vascular disease** |  |  |  |  |
|  | 9 (2.2%) | 46 (3.1%) | 355 (2.8%) | 410 (2.8%) |
| **Chronic obstructive lung disease** |  |  |  |  |
|  | 101 (24.9%) | 223 (14.9%) | 1516 (11.9%) | 1840 (12.6%) |
| **Dementia** |  |  |  |  |
|  | 4 (1.0%) | 1 (0.1%) | 47 (0.4%) | 52 (0.4%) |
| **Musculoskeletal disorders** |  |  |  |  |
|  | 11 (2.7%) | 22 (1.5%) | 197 (1.5%) | 230 (1.6%) |
| **Gastrointestinal disorders** |  |  |  |  |
|  | 8 (2.0%) | 15 (1.0%) | 109 (0.9%) | 132 (0.9%) |
| **Liver disease** |  |  |  |  |
| No | 395 (97.3%) | 1485 (99.1%) | 12598 (99.0%) | 14478 (99.0%) |
| Mild | 4 (1.0%) | 11 (0.7%) | 80 (0.6%) | 95 (0.6%) |
| Mod / severe | 7 (1.7%) | 3 (0.2%) | 45 (0.4%) | 55 (0.4%) |
| **Renal disease** |  |  |  |  |
|  | 17 (4.2%) | 61 (4.1%) | 543 (4.3%) | 621 (4.2%) |
| **Cancer** |  |  |  |  |
| No | 347 (85.5%) | 1387 (92.5%) | 11638 (91.5%) | 13372 (91.4%) |
| Non-metaststic | 34 (8.4%) | 75 (5.0%) | 733 (5.8%) | 842 (5.8%) |
| Metastatic | 25 (6.2%) | 37 (2.5%) | 352 (2.8%) | 414 (2.8%) |
| **Diabetes** |  |  |  |  |
| No | 352 (86.7%) | 1277 (85.2%) | 10713 (84.2%) | 12342 (84.4%) |
| Yes | 49 (12.1%) | 208 (13.9%) | 1813 (14.2%) | 2070 (14.2%) |
| With complications | 5 (1.2%) | 14 (0.9%) | 197 (1.5%) | 216 (1.5%) |
| **Cerebrovascular Disease** |  |  |  |  |
|  | 28 (6.9%) | 125 (8.3%) | 1030 (8.1%) | 1183 (8.1%) |
| **Charlson Index** |  |  |  |  |
| 0 | 136 (33.5%) | 340 (22.7%) | 5009 (39.4%) | 5485 (37.5%) |
| 1 | 117 (28.8%) | 620 (41.4%) | 3993 (31.4%) | 4730 (32.3%) |
| 2+ | 153 (37.7%) | 539 (36.0%) | 3721 (29.2%) | 4413 (30.2%) |
| **ACE-I** |  |  |  |  |
|  | 64 (15.8%) | 233 (15.5%) | 2378 (18.7%) | 2675 (18.3%) |
| **ARB** |  |  |  |  |
|  | 55 (13.5%) | 168 (11.2%) | 1456 (11.4%) | 1679 (11.5%) |
| **Statins** |  |  |  |  |
|  | 111 (27.3%) | 326 (21.7%) | 3373 (26.5%) | 3810 (26.0%) |
| **Aspirin** |  |  |  |  |
|  | 1 (0.2%) | 2 (0.1%) | 17 (0.1%) | 20 (0.1%) |
| **OAC admission** |  |  |  |  |
|  | 16 (3.9%) | 44 (2.9%) | 356 (2.8%) | 416 (2.8%) |
| **OAC on discharge** |  |  |  |  |
|  | 131 (32.3%) | 896 (59.8%) | 4340 (34.1%) | 5367 (36.7%) |
| **CHADS_VASC** |  |  |  |  |
| Mean (SD) | 2.27 (1.50) | 2.46 (1.45) | 2.22 (1.56) | 2.25 (1.55) |
| Median [Min, Max] | 2.00 [0, 7.00] | 2.00 [0, 7.00] | 2.00 [0, 9.00] | 2.00 [0, 9.00] |
| **Cardioversion** |  |  |  |  |
|  | 19 (4.7%) | 130 (8.7%) | 1166 (9.2%) | 1315 (9.0%) |

AMI = acute myocardial infarction, ACE-I = angiotensin converting enzyme inhibitors, ARB = angiotensin receptor blockers, OAC = oral anticoagulants

Table 2 shows the outcomes and median follow-up time according to discharge drug exposure. There was no statistically significant difference between the isolated beta blocker and digoxin groups for the composite outcome (risk difference (RD) 1.8%, 95% Confidence Interval (CI) -1.7 to 5.3, p = 0.3). The composite outcome was higher in the combined group compared to the beta blocker alone group (15.5% versus 11.5%, RD = 4.0%, 95% CI 2.0 to 5.9, p <0.001). This difference was largely secondary to more repeat CV hospitalizations in the combined group (14.4%) versus 10.7%, RD = 3.7, 95% CI 1.8 to 5.6, p < 0.001). There was no difference in hospitalizations between the isolated digoxin and beta blocker groups. The beta blocker group had the lowest mortality but with no statistically significant difference compared to the combined group (RD 0.3%, 95% CI -0.3 to 0.9, p = 0.3). Mortality in the digoxin alone group was increased compared to the beta blocker group (RD 1.9% 95% CI 0.2 to 3.6, p < 0.001)

**Table 2.** Outcome data (no adjustments for baseline confounders)

|  | Digoxin | BB + digoxin | BB | Overall |
| --- | --- | --- | --- | --- |
|  | (N=406) | (N=1499) | (N=12723) | (N=14628) |
| **Outcomes** |  |  |  |  |
| No outcomes | 352 (86.7%) | 1267 (84.5%) | 11259 (88.5%) | 12878 (88.0%) |
| CV hospitalization | 43 (10.6%) | 216 (14.4%) | 1366 (10.7%) | 1625 (11.1%) |
| Deaths | 11 (2.7%) | 16 (1.1%) | 98 (0.8%) | 125 (0.9%) |
| **Followup_time** |  |  |  |  |
| Mean (SD) | 404 (364) | 463 (390) | 468 (382) | 466 (382) |
| Median [Min, Max] | 286 [3.00, 1440] | 336 [3.00, 1450] | 358 [3.00, 1460] | 354 [3.00, 1460] |

Although not prespecified in our protocol, in a post hoc analysis, we examined individual CV hospitalization causes. The majority of the recurrent hospitalization were for atrial fibrillation and were less frequent in the beta blocker group compared to the combination group (995 (7.8%) versus 155 (10.3%), RD = 2.5%, 95% CI 0.1 - 4.2) but not different from the digoxin alone group (995 (7.8%) versus 32 (7.9%%), RD = 0.1%, 95% CI -0.1, 95% CI (-2.8, 2.7)). There were also fewer congestive heart failure admissions for the beta blocker alone group compared to the combined group (249 (2.0%) versus 48 (3.2%), RD 1.2%, 95% CI 0.3 - 2.2) but not compared to the digoxin alone group (249 (2.0%) versus 11 (2.7%), RD 0.7%, 95% CI -1.0 - 2.7). There was no statistically significant differences between the three groups for acute coronary syndrome hospitalizations (beta blocker alone 122 (0.9%) versus combined 13 (1.0%) versus digoxin alone 0, p = 0.1 for trend).

The Kaplan Meier survival curves for the composite endpoint is shown in Figure 1 (log rank test, p < 0.001) for this unadjusted analysis. However, given the important baseline imbalances observed in Table 1, adjusted time to event analyses are required. The baseline imbalances have been adjusted with propensity score inverse probability weighting and the success of this balancing procedure is shown in Figure 2 where the standardized mean differences are displayed and are judged acceptable (standardized mean differences < 0.1).

**Figure 1.**
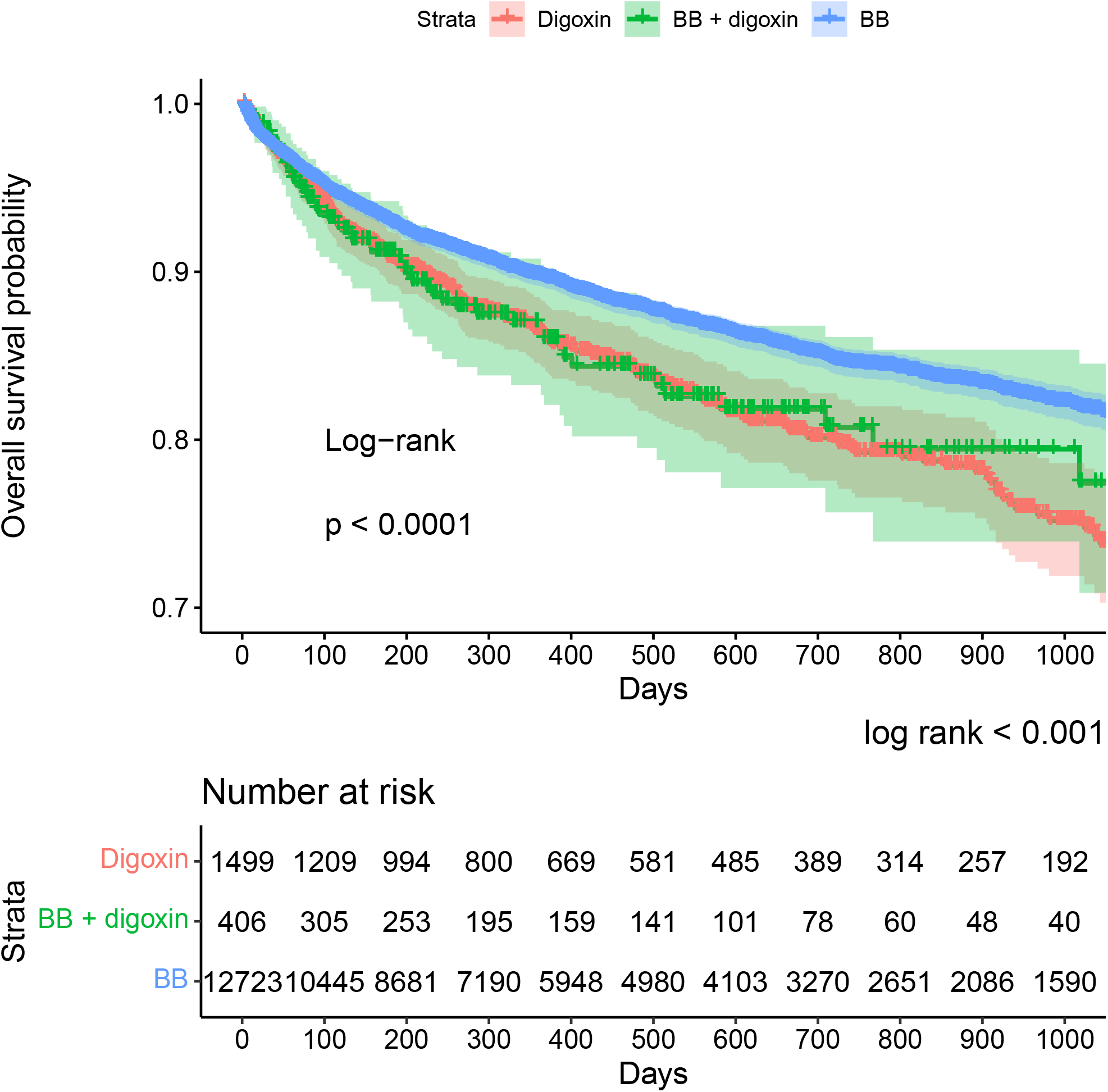
Kaplan Meier curve.

**Figure 2.**
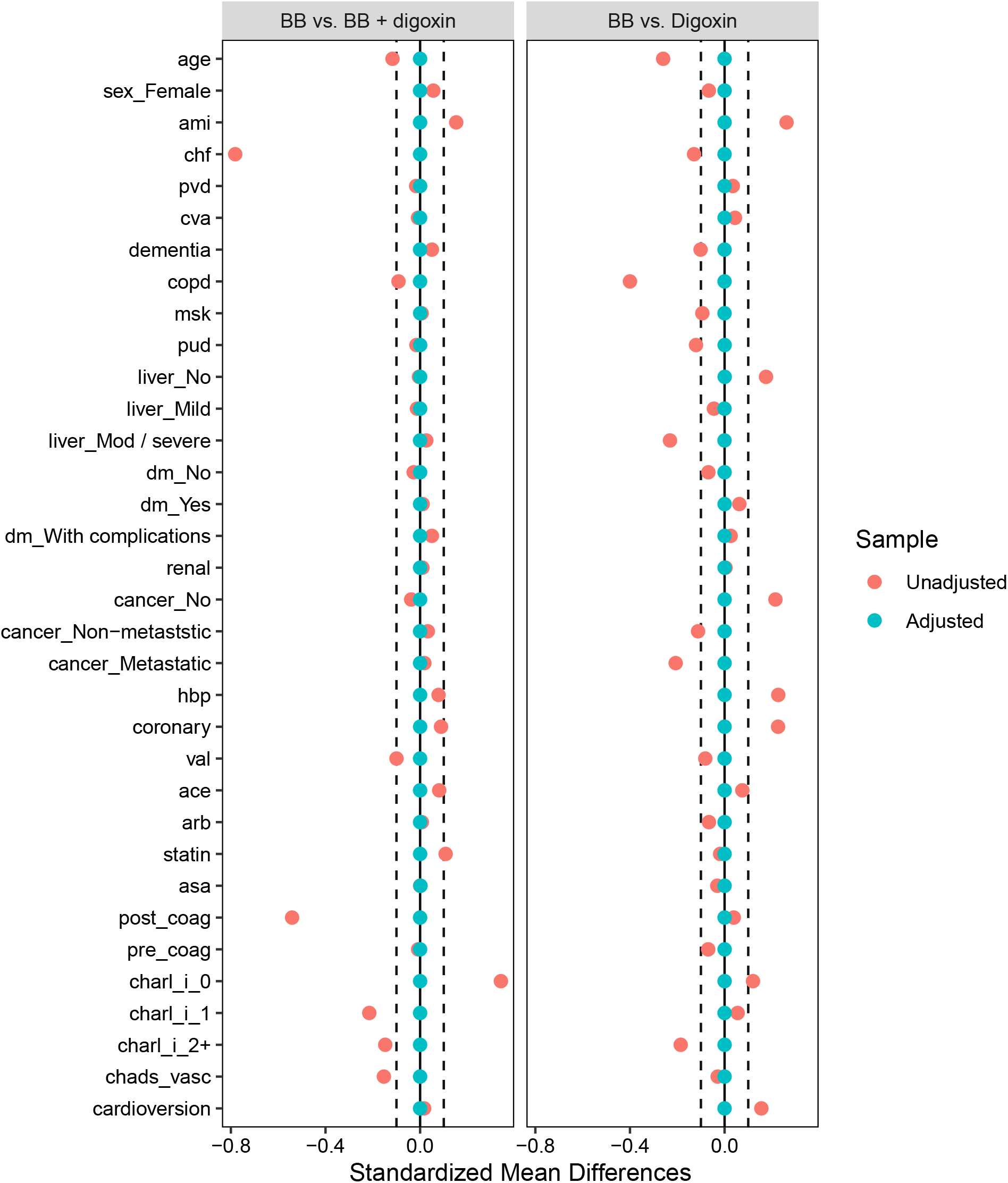
Standardized mean differences in raw and inverse probability weighted data. Figure 2 − Covariate balance before and after inverse probability weighting

The unadjusted and adjusted hazard ratios (aHR) from the Cox proportional models for the primary composite outcome with the matched dataset and the **ATT** pseudo population are displayed in Table 3. These results show that for this population of patients hospitalized for incident atrial fibrillation, compared to those discharged on isolated beta blocker therapy, patients additionally receiving digoxin (aHR 1.09, 95% CI 0.90 - 1.31) or receiving digoxin instead of beta blocker (aHR 1.24, 95% CI 0.85 - 1.81) did not have a statistically significant difference in the composite outcome of recurrent CV hospitalizations and death.

**Table 3.** Unadjusted and adjusted hzard ratios.

| Cox proportional hazard ratios |  |  |  |
| --- | --- | --- | --- |
| term | estimate | lower | higher |
| unadjusted model |  |  |  |
| BB (reference) | 1.00 | NA | NA |
| groupDigoxin | 1.28 | 0.98 | 1.68 |
| groupBB + digoxin | 1.36 | 1.18 | 1.56 |
| adjusted model |  |  |  |
| BB (reference) | 1.00 | NA | NA |
| groupDigoxin | 1.24 | 0.85 | 1.81 |
| groupBB + digoxin | 1.09 | 0.90 | 1.31 |

Sensitivity analyses employing a ATE estimand and constructing the propensity score model with a maximum likelihood algorithm produced identical results. The E-value, the minimum amount of unmeasured confounding needed to move the estimate and confidence interval for the combined group to include a 50% increase in risk compared to the beta blocker alone group is 2.1.

## 4. Discussion

In this cohort of patients with an incident hospitalization for atrial fibrillation, compared to those discharged on beta blockers alone there were no differences in the risk for recurrent CV hospitalizations or in-hospital death in patients who received a discharge prescription for digoxin in addition to, or in place of, a beta blocker prescription. These results were robust to different sensitivity analyses.

While RCTs remain the gold standard to evaluate drug efficacy, there are few contemporary trials that compare digoxin and beta blockers for long term clinical outcomes (mortality and repeat cardiovascular hospitalizations) for a patient population with an incident atrial fibrillation hospitalization. Previous meta-analyses of digoxin have largely been restricted to heart failure populations[18] but one previous meta-analysis[15] did examine the role of digoxin in atrial fibrillation and included 28 trials but only six trials reported results for mortality and none reported rehospitalization rates. The results from this later meta-analysis[15] are limited by the sample small size (digoxin 7 deaths in 269 vs, 10 10 deaths in 254, RR 0.82, 95% CI (0.34 - 2.00), multiple comparators (3 with amiodarone, 2 with verapamil and 1 with landiolol) and short follow-up times (<30 days). The only recent trial[12] comparing the effect of digoxin versus beta blocker (bisoprolol) in atrial fibrillation was again of modest size (160 subjects), concentrated on quality of life outcomes, and reported no differences. This trial had only 11 deaths (digoxin, 4 patients (5.0%) versus bisoprolol, 7 patients(8.8%), RR 0.73, 95% CI (0.24, 2.21)).

In this context, decisions must be informed from data from well performed non-experimental designs. Our study has several strengths including its incident design, thereby avoiding possible prevalence bias and limiting the study patient population to those prescribed beta-blockers, digoxin or their combination, in isolation of other anti-arrhythmic drugs, which should sharpen the exposure contrast. In addition, our study design attempted to emulate a target RCT and following these principles has been shown to minimize information, selection and immortal times biases[8],[9]. By following this approach and using propensity score methods to control for baseline confounding, potential biases from an observational study design may be reduced. Our propensity score methods suggest that measured confounding variables have been appropriately adjusted. Propensity score methods also separate the exposure and outcome models minimizing false positive errors from researcher degrees of freedom in the model selection process.

Our study does have limitations. First we do not know the exact reason for the choice of drug therapy and while our propensity score adjustments have successfully taken into consideration multiple baseline risk factors, there is no guarantee that these potential reasons, for example inability to control the ventricular rate, persistent symptoms, or drug intolerance have not influenced the outcomes. Similarly other unmeasured risk factors, including smoking, obesity, and socio-economic determinants, may be unequally distributed between the groups. However, in order for these non-measured variables to be confounders, physicians must be basing their choice of anti-arrhythmic drug on these characteristics, which seems somewhat unlikely. Moreover for the combined group, the unmeasured confounding necessary to move to statistically significant hazard ratio of 1.5 is substantial (EValue = 2.1). Another limitation is our use of an intention to treat paradigm where group assignment is determined at hospital discharge, and therefore exposure misclassification may occur over time if patients do not follow their initial treatments. Consequently, while these data sources have been used extensively in clinical research[14],[13], exposure, covariate and outcome misclassifications remain a possibility. This is perhaps especially true for mortality where the data set only records deaths that occur during a hospitalization. Also while we have more outcomes than what has been reported in previous studies[15], the sample size nevertheless lacks the statistical power to examine mortality as a separate outcome.

In conclusion, this study found that compared to isolated beta blocker therapy following a hospitalization for incident atrial fibrillation, the risk for the composite outcome of total hospital mortality or repeat cardiovascular hospitalizations was not increased by either the substitution or addition of digoxin. However, further studies to refine the precision of these findings would be helpful.

## Data Availability

All data produced in the present work are contained in the manuscript

